# Variants in *CALD1*, *ESRP1*, and *RBFOX1* are associated with orofacial cleft risk

**DOI:** 10.1101/2025.01.21.25320889

**Authors:** Jenna C. Carlson, Xinyi Zhang, Zeynep Erdogan-Yildirim, Terri H. Beaty, Azeez Butali, Carmen J. Buxó, Lord J.J. Gowans, Jacqueline T. Hecht, Ross E. Long, Lina Moreno, Jeffrey C. Murray, Ieda M. Orioli, Carmencita Padilla, George L. Wehby, Eleanor Feingold, Elizabeth J. Leslie-Clarkson, Seth M. Weinberg, Mary L. Marazita, John R. Shaffer

**Affiliations:** Department of Human Genetics, School of Public Health, University of Pittsburgh, Pittsburgh, PA; Department of Biostatistics and Health Data Science, School of Public Health, University of Pittsburgh, Pittsburgh, PA; Center for Craniofacial and Dental Genetics, Department of Oral and Craniofacial Sciences, School of Dental Medicine, University of Pittsburgh, Pittsburgh, PA; Department of Epidemiology, Johns Hopkins University, Baltimore, MD; Department of Oral Biology, Radiology, and Medicine, University of Iowa, Iowa City, IA; Dental and Craniofacial Genomics Core, School of Dental Medicine, University of Puerto Rico, San Juan, Puerto Rico; Department of Biochemistry and Biotechnology, Kwame Nkrumah University of Science and Technology, Kumasi, Ghana; Department of Pediatrics, McGovern Medical School University of Texas Health at Houston, Houston, TX; Lancaster Cleft Palate Clinic, Lancaster, PA; Department of Orthodontics & The Iowa Institute for Oral Health Research, University of Iowa, Iowa City, IA; Department of Pediatrics, University of Iowa, Iowa City, IA; INAGEMP (National Institute of Population Medical Genetics), Porto Alegre, Brazil; ECLAMC (Latin American Collaborative Study of Congenital Malformations) at Department of Genetics, Federal University of Rio de Janeiro, Rio de Janeiro, Brazil; Department of Pediatrics, College of Medicine, Institute of Human Genetics, National Institutes of Health, University of the Philippines Manila, Manila, The Philippines; Philippine Genome Center, University of the Philippines System, Quezon, The Philippines; Department of Health Management and Policy, College of Public Health, University of Iowa, Iowa, IA; Department of Statistics, Oregon State University, Corvallis, OR; Department of Human Genetics, Emory University School of Medicine, Atlanta, GA

## Abstract

Nonsyndromic orofacial clefts (OFCs) are common, heritable birth defects caused by both genetic and environmental risk factors. Despite the identification of many genetic loci harboring OFC-risk variants, there are many unknown genetic determinants of OFC. Furthermore, while the process of embryonic facial development is well characterized, the molecular mechanisms that underly it are not. This represents a major hurdle in understanding how disruptions in these biological processes result in OFC. Thus, we sought to identify novel OFC-risk loci through a genome-wide multi-ancestry study of five nested OFC phenotypes (isolated cleft lip [CLO], isolated cleft palate [CPO], cleft lip and palate [CLP], cleft lip with/without cleft palate [CL/P], and any cleft [ANY]) representing distinct cleft subtypes to identify subtype-specific signals and grouped types to maximize power to detect shared genetic effects. We performed genome-wide meta-analyses of these five OFC phenotypes from three cohorts totaling >14,000 individuals using METAL. In addition to replicating 13 known OFC-risk loci, we observed novel association in three regions: the 1p36.32 locus (lead variant rs584402, an intergenic variant, p_CLO_ = 3.14e-8), the 7q33 locus (lead variant rs17168118, an intronic variant in *CALD1*, p_CLP_ = 9.17e-9), and the 16p13.3 locus (lead variant rs77075754, an intronic variant in *RBFOX1*, p_CL/P_ = 1.53e-9, p_ANY_ = 1.93e-9). We also observed a novel association within the known risk locus 8q22.1 that was independent of the previously reported signal (lead variant rs4735314, an intronic variant in *ESRP1*, p_CLP_ = 1.07e-9, p_CL/P_ = 3.88e-8). Next, we performed multi-tissue TWAS with s-MulTiXcan and identified four overlapping genes with significant genetically predicted transcription associated with OFC risk. These genes also overlapped the genome-wide significant association signals from the meta-analysis, including *CALD1* and *ESRP1* and known OFC-risk genes *TANC2* and *NTN1*. Each of the newly reported loci has potential regulatory effects, including evidence of craniofacial enhancer activity, that offer new clues as to the molecule mechanisms underlying embryonic facial development.

**Author Summary:** Orofacial clefts, including cleft lip and cleft palate, are common birth defects that can be caused by both genetic and environmental factors. While many of these factors are known, there are still significant gaps in our understanding of how and why clefts arise. To help address this deficiency, we measured association between variants across the genome and clefting in cases and controls from diverse genetic ancestries. We identified three new candidate genes (*CALD1*, *ESRP1*, and *RBFOX1*) that may be involved in cleft risk and reaffirmed the role of 12 previously reported risk genes. We also found evidence that clefting was associated with predicted gene expression at *CALD1* and *ESRP1* and that the associated variants in these genes were located near regions known to be involved in the regulation of gene expression in craniofacial tissues during development.

## Introduction

Orofacial clefts (OFC) are common, heritable birth defects caused by both genetic and environmental risk factors(1). OFCs have a heterogeneous presentation and can occur in isolation (isolated) or simultaneously with other birth defects (non-isolated)(1–4). Isolated OFCs, which affect ∼1/700 births worldwide, consist of clefts of the lip and/or palate. OFCs arise through disruption of the formation of the lip and/or palate, which involves a complex series of events spanning weeks 4-10 of human development(2). While this process is well characterized, the underlying molecular mechanisms that lead to clefting are not. Knowledge of the various genes and pathways involved is critical to this endeavor.

The genetic etiology of OFCs is complex, characterized by both allelic and locus heterogeneity (5). Moreover, the three main forms of nonsyndromic OFC (cleft lip only or CLO, cleft palate only or CPO, and cleft lip and palate or CLP) are believed to have both shared and subtype-specific genetic risk factors(1,3,4). Despite the identification of over 60 genetic loci harboring OFC-risk variants(6), these only account for an estimated 25% of the heritability of isolated OFC(7), suggesting that there are more genetic determinants yet to be discovered. Moreover, the biological interpretation of these loci is challenging since most OFC-risk variants fall outside protein-coding gene regions, making their functional impact a challenge to decipher.

In addition to differences across cleft subtypes, there are large differences in OFC prevalence according to genetic ancestry (8,9). For example, OFC risk is higher in individuals of recent East Asian ancestry compared to individuals of recent European or Sub-Saharan African ancestry. Such differences make it essential to include diverse cohorts in our genetic analyses, yet most studies of OFC inadequately address this concern.

In the current study, we attempt to address some of these gaps by conducting genome-wide meta-analyses of three ancestrally diverse cohorts of individuals and trios with isolated OFCs (hereafter simply referred to as OFCs).

## Results

We performed genome-wide meta-analyses from three studies totaling 14,481 individuals from diverse genetic ancestries for five cleft phenotypes: CLO, CPO, CLP, cleft lip with or without cleft palate (CL/P), and any cleft (ANY) (**S1-S5 Figures**). We observed genome-wide significant (p < 5e-8) signals at 16 loci (**Table 1**), including 13 known OFC-risk loci: *PAX7* (1p36.3), *ARHGAP29* (1p22.1), *IRF6* (1q32.2), *THADA* (2p21), *FGF10* (5p12), 8q21.3, 8q22.1, 8q24.21, *VAX1* (10q25.3), *SPRY2* (13q31.1), *NTN1* (17p13.1), *TANC2* (17q23.2), and *MAFB* (20q12). At some of these loci, there were multiple independent signals, including two at *PAX7*, five at *IRF6*, five at 8q24.21, three at *VAX1*, two at *SPRY2*, and two at *NTN1* (17p13.1) (**Table 1**).

**Table 1.** Results from genome-wide meta-analysis for each independent lead variant demonstrating genome-wide significance (p < 5e-8) in at least one phenotype. Columns include Chromosome (Chr), position (hg38), reference and alternate (effect) alleles (Ref/Alt), rsID, candidate gene (either the gene nearest the variant or a previously implicated risk gene), direction of effect (Dir) on any cleft for POFC1, POFC2, and GENEVA OFC, respectively (? indicates variant was not present in that study), and p-values and from meta-analysis across each phenotype definition.

| Region | Chr | Pos | Ref | Alt | rsID | Candidate Gene | Dir | P ANY | P CL/P | P CLP | P CLO | P CPO |
| --- | --- | --- | --- | --- | --- | --- | --- | --- | --- | --- | --- | --- |
| **1p36.32** | **1** | **5238225** | **C** | **T** | **rs584402** | **AJAP1** | **++-** | **4.74E-03** | **1.02E-03** | **7.09E-02** | **3.14E-08** | **4.81E-01** |
| 1p36.13 | 1 | 18645140 | G | C | rs56675509 | PAX7 | +++ | 6.84E-10 | 1.41E-12 | 2.08E-09 | 1.67E-07 | 8.53E-01 |
| 1p36.13 | 1 | 18646288 | TTC | T | rs10546636 | PAX7 | +++ | 1.11E-10 | 1.11E-13 | 4.43E-11 | 2.18E-07 | 3.47E-01 |
| 1p22.1 | 1 | 94092554 | G | T | rs66515264 | ARHGAP29 | ++? | 6.46E-09 | 3.30E-10 | 2.82E-08 | 1.94E-05 | 2.11E-01 |
| 1q32.2 | 1 | 209846668 | C | A | rs3766612 | IRF6 | +++ | 5.50E-10 | 9.61E-12 | 1.71E-05 | 2.19E-12 | 6.07E-01 |
| 1q32.2 | 1 | 209942373 | C | A | rs201670404 | IRF6 | --- | 2.00E-08 | 7.63E-11 | 2.56E-09 | 6.16E-05 | 4.22E-01 |
| 1q32.2 | 1 | 209798853 | G | A | rs4844494 | IRF6 | --- | 3.95E-18 | 1.00E-22 | 1.74E-16 | 3.13E-12 | 1.29E-01 |
| 1q32.2 | 1 | 209815644 | G | A | rs17015268 | IRF6 | --- | 4.30E-18 | 5.42E-23 | 1.31E-16 | 1.05E-12 | 8.93E-02 |
| 1q32.2 | 1 | 209815936 | G | A | rs77542756 | IRF6 | --- | 4.12E-18 | 7.23E-23 | 1.74E-16 | 9.96E-13 | 9.63E-02 |
| 2p21 | 2 | 43286897 | G | A | rs11900952 | THADA | --- | 3.24E-08 | 5.83E-09 | 1.44E-07 | 2.21E-05 | 3.35E-01 |
| 5p12 | 5 | 44263295 | C | T | rs6883600 | FGF10 | ++? | 4.21E-07 | 4.23E-08 | 1.40E-05 | 3.12E-07 | 4.57E-01 |
| **7q33** | **7** | **134892089** | **A** | **G** | **rs17168118** | **CALD1** | **++?** | **1.44E-06** | **5.77E-08** | **9.17E-09** | **2.81E-03** | **8.55E-01** |
| 8q21.3 | 8 | 87892130 | C | T | rs11786656 | DCAF4L2 | --- | 8.98E-09 | 1.94E-11 | 3.03E-09 | 6.68E-06 | 5.92E-01 |
| **8q22.1** | **8** | **94656529** | **T** | **C** | **rs4735314** | **ESRP1** | **---** | **7.46E-06** | **3.88E-08** | **1.07E-09** | **1.53E-02** | **2.82E-01** |
| 8q24.21 | 8 | 128952627 | G | A | rs17242358 | CCDC26 | +++ | 2.37E-27 | 4.14E-35 | 9.74E-25 | 2.45E-19 | 4.41E-01 |
| 8q24.21 | 8 | 128754576 | A | G | rs2349688 | CCDC26 | --- | 4.31E-08 | 2.68E-10 | 9.18E-08 | 2.23E-06 | NA |
| 8q24.21 | 8 | 128965218 | T | C | rs1470206 | CCDC26 | --- | 1.34E-13 | 1.34E-15 | 2.16E-10 | 2.56E-08 | 9.10E-01 |
| 8q24.21 | 8 | 128886123 | A | T | rs2119756 | CCDC26 | ??- | 9.49E-05 | 4.49E-08 | 1.91E-03 | 4.67E-07 | 6.37E-02 |
| 8q24.21 | 8 | 128969112 | C | T | rs11997787 | CCDC26 | +++ | 1.03E-14 | 4.40E-18 | 3.06E-12 | 7.41E-09 | 4.46E-01 |
| 10q25.3 | 10 | 117076565 | AC | A | rs5788208 | VAX1 | +++ | 1.42E-08 | 1.24E-09 | 5.33E-08 | 5.90E-05 | 8.73E-01 |
| 10q25.3 | 10 | 117081903 | G | T | rs11197889 | VAX1 | +++ | 9.14E-09 | 9.15E-10 | 5.00E-08 | 4.73E-05 | 9.17E-01 |
| 10q25.3 | 10 | 117081904 | C | T | rs11197890 | VAX1 | +++ | 7.67E-09 | 8.08E-10 | 5.11E-08 | 4.16E-05 | 9.29E-01 |
| 13q31.1 | 13 | 80126941 | G | C | rs11842594 | SPRY2 | +++ | 2.50E-07 | 1.51E-09 | 1.14E-10 | 3.93E-02 | 5.57E-01 |
| 13q31.1 | 13 | 80127350 | T | C | rs1854110 | SPRY2 | --- | 1.49E-07 | 1.38E-09 | 1.54E-10 | 3.53E-02 | 6.62E-01 |
| **16p13.3** | **16** | **6429186** | **A** | **G** | **rs77075754** | **RBFOX1** | **??+** | **1.93E-09** | **1.53E-09** | **1.31E-06** | **2.36E-04** | **NA** |
| 17p13.1 | 17 | 9026902 | AACCCAAAACC<br>CAC | A | rs11273201 | NTN1 | --- | 2.19E-12 | 4.22E-14 | 2.42E-13 | 3.71E-05 | 5.19E-01 |
| 17p13.1 | 17 | 9017282 | T | C | rs4791330 | NTN1 | --- | 1.39E-08 | 4.72E-11 | 1.90E-12 | 1.25E-02 | 4.23E-01 |
| 17q23.2 | 17 | 62971963 | C | T | rs72843143 | TANC2 | --- | 6.04E-05 | 3.69E-08 | 5.78E-06 | 5.33E-05 | 1.18E-01 |
| 20q12 | 20 | 40613502 | G | C | rs11700188 | MAFB | ??- | 1.98E-10 | 1.20E-11 | 4.56E-11 | 2.25E-02 | 2.76E-01 |

Abbreviations ANY: any cleft, CL/P: cleft lip with/without cleft palate, CLP: cleft lip and palate, CLO: cleft lip only, CPO: cleft palate only. Bold values indicate novel associations

We observed four novel associations at biologically plausible loci at 8q22.1, 1p36.32, 7q33, and 16p13.3. At the 8q22.1 locus, we observed an association between rs4735314 (8:94656529 T>C) with CLP and CL/P (p_CLP_ = 1.07e-9, p_CL/P_ = 3.88e-8). While the 8q22.1 locus has been implicated in prior studies, the signal we observed at this region was independent from what has been previously reported (10) and contained seven intronic variants in *ESRP1* (**Figure 1A**). The previously identified variants at this locus (rs12681366 and rs957448) were 267 and 127 kb away, respectively, from the signal observed here and were not in linkage disequilibrium (r^2^ < 0.2) with associated variants in this study. In addition, we also observed association signals at three novel loci, 1p36.32, 7q33, and 16p13.3. The 1p36.32 locus was associated with CLO (rs584402, chr1:5238225 C>T, p_CLO_ = 3.17e-8) and the 95% credible set at this locus included the lead variant (rs584402) and six additional variants (rs586165, chr1:5237827 T>C, p_CLO_ = 6.76e-8; rs679712, chr1:5238707 T>A, p_CLO_ = 4.77e-8; rs551536, chr1:5238999 T>C, p_CLO_ = 5.84e-8; rs551376, chr1:5239052 G>A, p_CLO_ = 4.77e-8; rs549773, chr1:5239163 C>A, p_CLO_ = 3.45e-8; rs5772197, chr1:5239601 ACT>A, p_CLO_ =3.14e-8) which are all intergenic variants ∼ 446 kb downstream of *AJAP1* (**Figure 1B**). The 7q33 locus was associated with CLP (lead variant rs17168118, chr7:134892089 A>G, p_CLP_ = 9.16e-9) and the 95% credible set at this locus included the lead variant (rs17168118) and three additional variants (rs2075463, chr7:134867451 G>A, p_CLP_ = 7.48e-7; rs58715694, chr7:134874323 G>C, p_CLP_ = 6.17e-7; rs10488465, chr7:134877068 T>C, p_CLP_ = 2.29e-7), all intronic variants in *CALD1* (**Figure 1C**). The third novel locus, at 16p13.3, was associated with CL/P and any cleft (lead variant rs77075754, chr16: 6429186 A>G, p_CL/P_ = 1.52e-9, p_ANY_ = 1.92e-9), which was driven by the GENEVA OFC cohort. The three additional SNPs that comprise the 95% credible set in this region (rs79685543, chr16:6430284 C>A, p_CL/P_ = 2.11e-9; rs75282824, chr16:6439230 A>AT, p_CL/P_ = 1.11e-8; rs150962530, chr16:6440126 G>A, p_CL/P_ = 3.42e-9) are all contained within intron 2 of the MANE transcript of *RBFOX1* (**Figure 1D**). The association at this locus was only observed in the GENEVA OFC cohort as the minor allele frequency (MAF) was below the 0.05 threshold used in analyses of the other two cohorts. The direct examination of the lead SNP rs77075754 did not show evidence of association with CL/P in POFC1 (p_CL/P_ = 0.71, MAF = 0.013) or POFC2 (p_CL/P_ = 0.83, MAF = 0.033).

**Figure 1.**
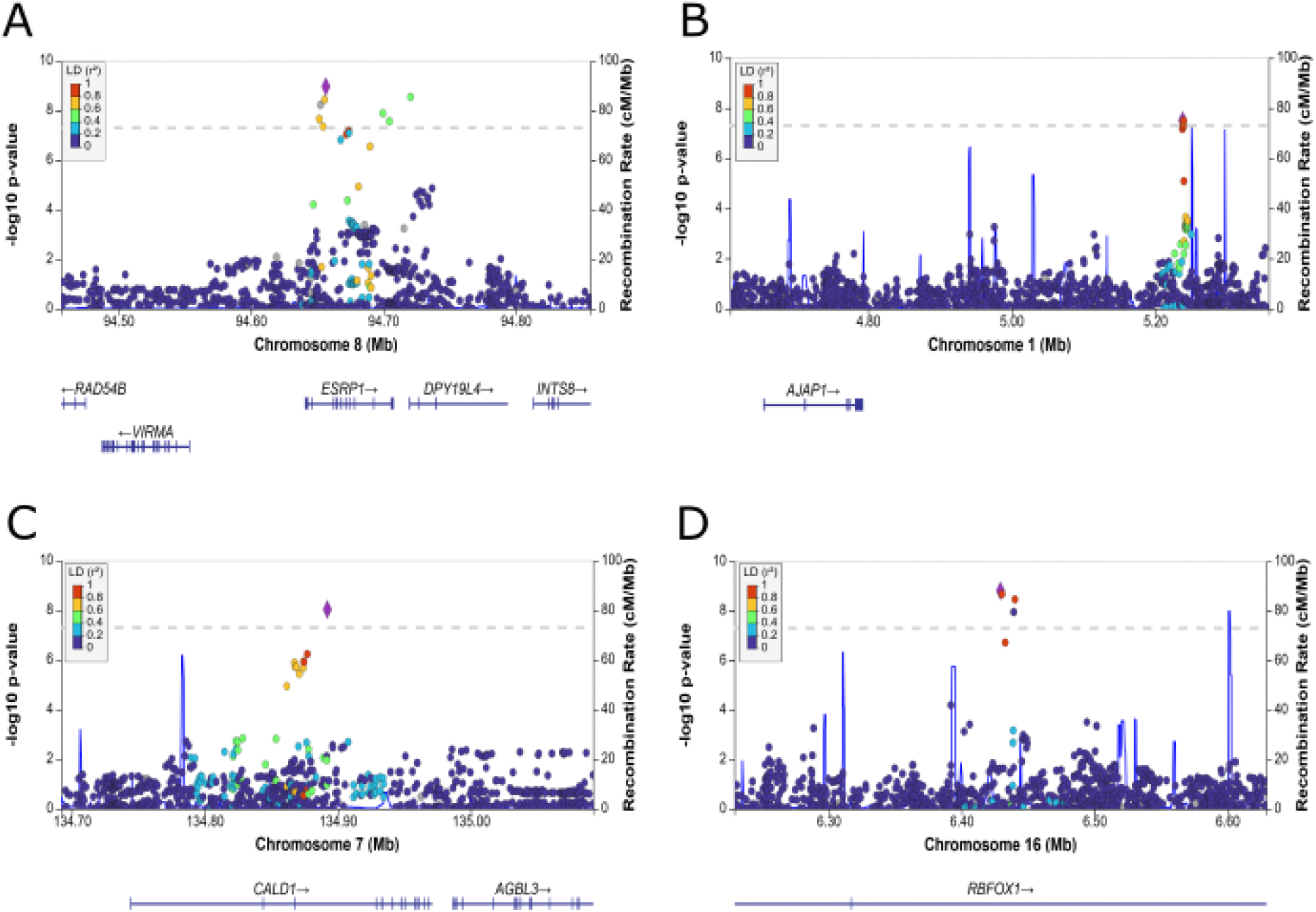
Regional association plots of meta-analyses for (A) 8q22.1 with cleft lip and palate (CLP), (B) 1p36.32 with CLO, (C) 7q33 with CLP, and (D) 16p13.3 with cleft lip with/without cleft palate (CL/P). Association results were obtained via fixed effects p-value based meta-analysis using METAL (44). Variants are colored based on LD with the top variant in each region (shown in purple) based on the 1000 Genomes Phase 3 all populations references. Positions are in hg38. Plots were generated using LocusZoom (45).

We also examined predicted gene expression to identify genetic regulatory mechanisms associated with orofacial clefting. For each phenotype, we also performed multi-tissue transcriptome-wide association studies (TWAS, **S6-S10 Figures**) and examined the overlap with loci discovered in the genome-wide meta-analyses. We observed significant associations between genetically predicted expression and OFC risk for at least one OFC phenotype, implicating a total of 512 genes (**S2 Table; Figure 2**). Of these, four genes overlapped with a genome-wide significant signal from the meta-analysis and demonstrated significant association with genetically predicted expression and more than one OFC trait: *NTN1* (p_ANY_ = 1.66e-14, p_CL/P_ = 9.36e-20, p_CLP_ = 8.08e-25), *CALD1* (p_ANY_ = 2.925e-12, p_CL/P_ = 1.26e-15, p_CLP_ = 3.61e-16), *ESRP1* (p_ANY_ = 4.95e-8, p_CL/P_ = 7.92e-12, p_CLP_ = 6.89e-11)*, and TANC2* (p_CL/P_ = 1.27e-10, p_CLP_ = 2.24e-8). We also performed gene set enrichment analysis of the 512 significant genes to identify significantly enriched biological processes, cellular components, and molecular function. We identified 10 significantly enriched biological processes including those relating to growth and development (GO:0040008, GO:0040007, GO:0048731) and response to stimuli (GO:0070887, GO:0050896) and one cellular component (cell periphery, GO:0071944) (**Table 2**).

**Figure 2.**
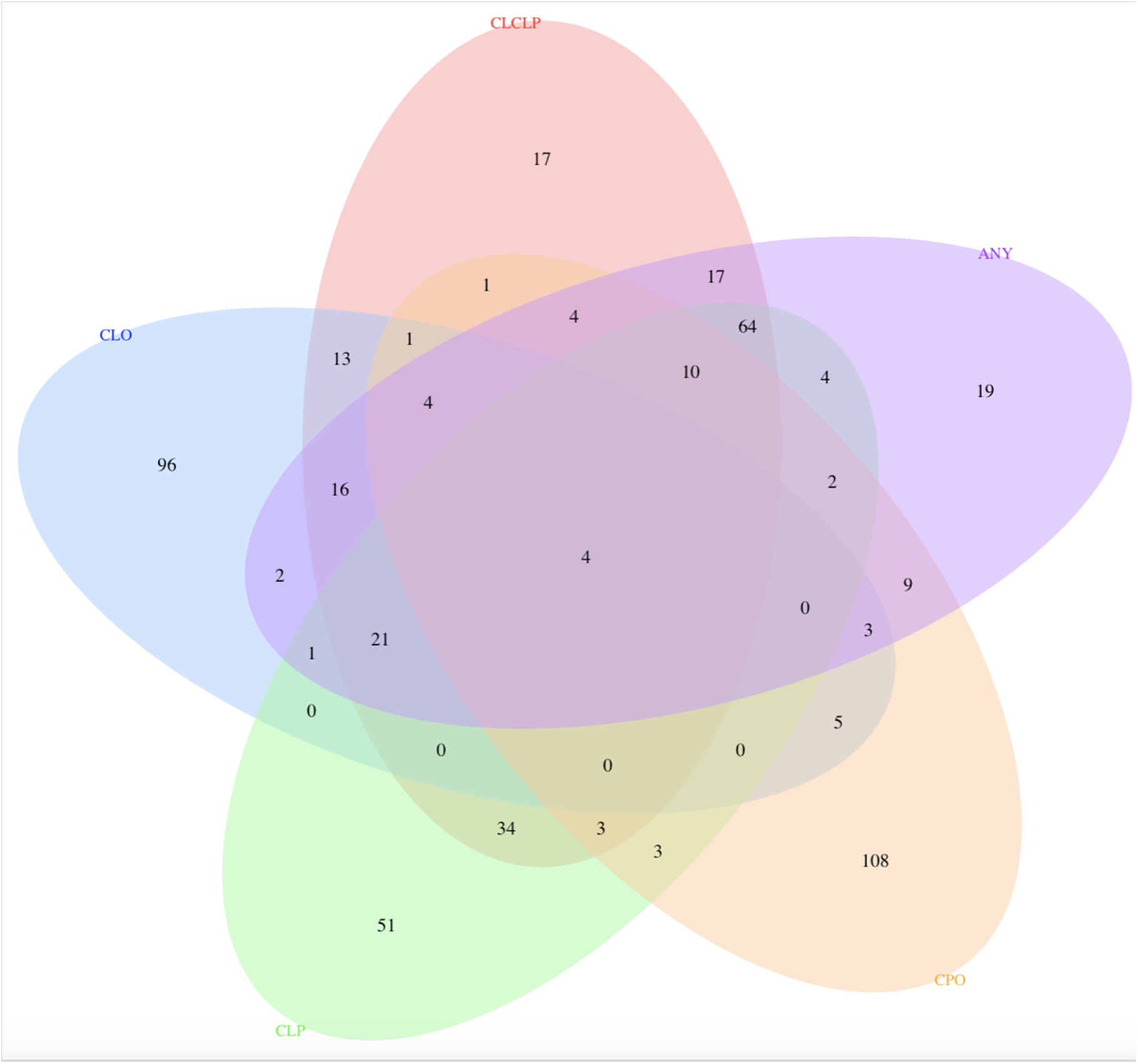
Venn Diagram showing the number of genes significant in TWAS results for the five cleft types. Colors indicate cleft type definition: ANY in purple, CL/P in orange, CLP in green, CLO in blue, and CPO in yellow. There were four genes associated with each cleft type definition (PTPRS, C9orf170, RP1-40E16.9, RP11-87G24.6).

**Table 2.** Significantly enriched Gene Ontology (GO) terms among the 512 genes showing significant results in the TWAS. Enrichment analyses were conducted using the multi-query in g:Profiler (11). Columns include source, term name, GO id, and adjusted p-values across each phenotype definition. P-values were adjusted to maintain an experiment-wide type 1 error rate of 0.05 using the g:SCS algorithm which accounts for the dependent structure of functionally annotated gene sets.

| Source | Term name | Term id | adj p ANY | adj p CL/P | adj p CLP | adj p CLO | adj p CPO |
| --- | --- | --- | --- | --- | --- | --- | --- |
| GO:BP | multicellular organismal process | GO:0032501 | 0.00008 | 0.00001 | 0.00001 | 1 | 1 |
| GO:BP | cellular response to chemical stimulus | GO:0070887 | 1 | 0.09717 | 0.00006 | 1 | 1 |
| GO:BP | regulation of growth | GO:0040008 | 0.00341 | 0.00138 | 0.06788 | 1 | 1 |
| GO:BP | growth | GO:0040007 | 0.03217 | 0.00180 | 0.01245 | 1 | 1 |
| GO:BP | positive regulation of sarcomere organization | GO:0060298 | 1 | 1 | 1 | 1 | 0.00242 |
| GO:BP | response to stimulus | GO:0050896 | 0.00624 | 0.00960 | 0.25129 | 1 | 1 |
| GO:BP | system development | GO:0048731 | 0.00914 | 0.00788 | 0.00937 | 1 | 1 |
| GO:BP | regulation of biological quality | GO:0065008 | 1 | 0.14047 | 0.01793 | 1 | 1 |
| GO:BP | thyroid hormone generation | GO:0006590 | 1 | 1 | 0.02126 | 1 | 1 |
| GO:BP | formation of primary germ layer | GO:0001704 | 1 | 0.04015 | 1 | 1 | 1 |
| GO:CC | cell periphery | GO:0001704 | 0.02245 | 0.00003 | 0.06350 | 0.04255 | 1 |

Abbreviations GO: gene ontology, BP: biological process, CC: cellular component, ANY: any cleft, CL/P: cleft lip with/without cleft palate, CLP: cleft lip and palate, CLO: cleft lip only, CPO: cleft palate only

## Discussion

Through meta-analysis of >14,000 individuals and subsequent TWAS, we identified four novel loci associated with risk of nonsyndromic orofacial clefting on 1p36.32, 7q33, and 16p13.3, each pointing to biologically relevant candidate genes that play a role in embryonic craniofacial development (discussed below). We also identified a new independent signal and candidate gene, *ESRP1*, near a previously associated locus at 8q22.1.

The association signal on 1p36.32 was uniquely seen in the GWAS for CLO and falls in a large intergenic region ∼446 Kb downstream of *AJAP1* and ∼753 upstream of *KCNAB2* and is near a known enhancer element (GH01J005235) of *AJAP1* and *KCNAB2* (12). *AJAP1* is a gene involved in the negative regulation of cell-matrix adhesion and wound healing (13) and is downregulated in human craniofacial tissues during CS14-22 compared to CS13 and control cells (14). *KCNAB2* is a known gene involved in 1p36 deletion syndrome, which is characterized by distinct craniofacial features including microbrachycephaly and can include orofacial clefts (15,16). This novel association may highlight the regulatory role of this region (either through *AJAP1*, *KCNAB2*, or both) in the development of cleft lip.

The 7q33 locus is spanned by *CALD1*, a gene encoding a calmodulin- and actin-binding protein that plays an essential role in the regulation of smooth muscle and nonmuscle contraction (13). Preclinical models suggest Cald1 has a role in building morphology through blood vessel endothelial cell migration and tube morphogenesis(17–20). CALD1 is also highly expressed in human and murine craniofacial tissue across CS13-20/E8.5-10.5, during which the formation of the lip and palate occurs (14,21). The association signal in this region is led by a cross-tissue eQTL for CALD1 and directly overlaps a known craniofacial super-enhancer as well as 6 other super-enhancers (aorta, duodenum smooth muscle, osteoblast, myotube, stomach smooth muscle and u87 cells) affecting CALD1 expression(22). Together, this suggests that CALD1 plays an important role in embryonic craniofacial development.

The signal on 16p13.3 is contained between two recombination hotspots located within a much larger topologically associated domain spanning the first two introns of *RBFOX1*. *RBFOX1* encodes an RNA binding protein and regulates alternative splicing of transcripts(13). In humans, RBFOX1 is expressed in neurons, heart, and muscle (23) as well as within muscle precursor cells from human craniofacial tissue (14). RBFOX1 is highly conserved and predicted intolerant to loss of function (loeuf 0.31)(24). In mice, Rbfox1 was differentially expressed during neural tube development (downregulated at E9.5, during the fusion of the neural tube)(25). However, some caution should be taken in this interpretation, as the association at this locus was only observed in the GENEVA OFC cohort and replication is still needed for this locus.

Association with orofacial clefting at the 8q22.1 locus has been previously reported for two SNPs: rs957448 (chr8:94529074 A>G) an intronic SNP in *VIRMA*, and rs12681366 (chr8: 94389037 T>C) an intronic SNP in *RAD54B* (10). In our analysis, we identified a separate signal led by rs4735314 not in LD with these two previously reported variants (LD r^2^ < 0.2). This new signal entirely comprises intronic SNPs in *ESRP1*, a gene that was also nominated through the multi-tissue TWAS. Interestingly, each of these three variants are significant eQTLs for both ESRP1 and a divergent transcript of VIRMA (VIRMA-DT, ENSG00000253704.3) in some tissues (23). *ESRP1* and its paralog *ESRP2* are epithelial cell-type-specific splicing regulators that regulate craniofacial development and the splicing of craniofacial and cleft associated genes including *CTNND1* and the FGFR2 signaling pathway (13,26,27). Mutations in *ESRP2* and *ESRP1/2* splicing target *CTNND1* are associated with orofacial clefts (27,28). In addition, mutations in *FGFR2* and other FGF genes lead to craniosynostosis, syndactyly, and cleft palate (29–33), demonstrating the potential importance of *ESRP1* to cleft risk. ESRP1 and VIRMA are targets for known super enhancers (from colon crypt tissue, gastric tissue, NHEK cells, HCC1954 cells, ACO 9m cells) as well as craniofacial specific enhancers (12,22). Within human craniofacial tissue, ESRP1 is solely expressed in ectodermal cells, and in bulk RNA-seq shows differential expression at critical stages for facial formation (CS13, CS14, CS15, CS17, and CS20) (14). RAD54B, the gene suggested by a previous study (34,35), is also differentially expressed in human craniofacial tissue across the period critical to facial development like ESRP1, but is not cell-sub-type-specific in its expression (14). VIRMA, another candidate gene in the region, is expressed in human craniofacial tissue but is not differentially expressed during the critical period of lip and palate formation (14). Together, this evidence supports *ESRP1* as a strong orofacial cleft risk locus warranting further functional investigation.

Through TWAS, we identified numerous genes with potential regulatory roles in OFC risk, many overlapping with GWAS signals. These TWAS results were enriched for many biological processes including those involved in growth and response to stimuli. These findings reflect that OFC risk is genetically regulated through several mechanisms—both those that directly regulate embryonic growth and those that respond to certain in-utero environmental exposures. The TWAS results additionally supported the potential regulatory role of *CALD1* and *ESRP1* in association with clefting as well as two known genes associated with OFC-risk, *NTN1* and *TANC2*.

As expected, nearly every locus we identified demonstrated genome-wide significance with CL/P, recapitulating the evidence for shared genetic architecture between CLP and CLO for many loci. However, this was not the case for the novel associations with the *AJAP1/KCNAB2* (1p36.32) locus, which was only seen in CLO alone, and with *CALD1*, which only surpassed genome-wide significance when examining CLP alone, and not combining it with CLO as a composite phenotype as is typically done to maximize statistical power. While the *CALD1* locus still demonstrated association with the composite phenotype CL/P, this example highlights the importance of examining subtype-specific associations to maximize power to detect loci enriched for association within one phenotype definition. None of the loci observed in this study were associated with CPO, despite prior evidence of association with *GRHL3* in a subset of the individuals used in this meta-analysis, all of European genetic ancestry (36).

In summary, this study provides strong evidence for two novel genes (*CALD1* at 7q33 and *ESRP1* at 8q22.1), in association with orofacial cleft risk through transcriptomic and genomic analyses. These loci are each associated with many orofacial cleft phenotypes (CLO, CLP, CL/P, and/or any cleft) and are regulated by both craniofacial-specific and cross-tissue super enhancers, suggesting that they may represent more global regulators of embryonic growth and development beyond their role in facial formation. This study also demonstrates the utility of multi-tissue TWAS and colocalization for identifying these putative regulators, despite the absence of a relevant tissue type in existing eQTL databases.

## Methods

### Samples and Genotyping

Samples and data for these analyses were derived from three studies of orofacial clefts and are summarized in **Table 3**. The first, referred to here as GENEVA OFC, comprised 5,856 individuals from 1,953 case-parent trios of isolated, nonsyndromic orofacial clefts that were recruited from several sites across the United States, Western Europe (Norway, Denmark), and East Asia (Korea, Singapore, Taiwan, Philippines, and China) as part of the International Consortium to Identify Genes and Interactions Controlling Oral Clefts, part of the Gene Environment Association Studies (GENEVA) initiative (dbGaP accession number phs000094.v1.p1). More details about recruiting these participants and genotyping QC have been described elsewhere(37). Briefly, samples were genotyped for 589,945 SNPs on the Illumina Human610-Quadv.1_B BeadChip. A total of 412 overlapping individuals who were re-genotyped in subsequent studies were excluded from the analysis of GENEVA OFC herein.

**Table 3.** Sample size by phenotype.

|  | Controls | CLO | CPO | CLP | CL/P | ANY <sup>a</sup> |
| --- | --- | --- | --- | --- | --- | --- |
| GENEVA OFC (# trios) | --- | 431 | 455 | 1064 | 1495 | 1953 |
| POFC1 | 2620 | 624 | 299 | 1946 | 2570 | 2881 |
| POFC2 | 1503 | 469 | 172 | 980 | 1449 | 1621 |

<sup>a</sup> includes some cases with unknown cleft sub-type
Abbreviations ANY: any cleft, CL/P: cleft lip with/without cleft palate, CLP: cleft lip and palate, CLO: cleft lip only, CPO: cleft palate only

The second study, referred to here as POFC1, included 2,881 cases and 2,620 controls (unaffected individuals from families with no reported history of OFCs) from the Genetics of Orofacial Clefts and Related Phenotypes study (dbGaP accession number phs000774.v2.p1). Participants were recruited from 13 countries in North America (United States), Central or South America (Guatemala, Argentina, Colombia, Puerto Rico), Asia (China, Philippines), Europe (Denmark, Turkey, Spain), and Africa (Ethiopia, Nigeria). Samples were genotyped for 539,473 SNPs on the Illumina HumanCore+Exome array. Additional details on recruitment, genotyping, and quality controls have been previously described.(36,38)

The third study, referred to here as POFC2, included 1,621 cases and 1,503 controls from the Genetics of Orofacial Clefts, Sub-types, and Subclinical Phenotypes study (dbGaP accession number phs002815.v2.p1). Genotyping was performed at CIDR using the Infinium Global Diversity Array-8 v1.0 Array along with extensive quality control following the methods of Laurie et al.(39) yielding 1,434,549 SNPs.

Written informed consent was obtained for all participants and all sites had both local IRB approval and approval at the University of Pittsburgh, the University of Iowa, or Johns Hopkins University.

For each study, we imputed genotypes with the TOPMed reference panel via the TOPMed Imputation Server(40). We removed variants with poor imputation quality (r^2^ < 0.80) and extracted the most likely genotype, keeping only those with a minimum genotype probability of 0.9. Prior to association testing we also removed variants with Mendelian error rates >0.5% [in GENEVA OFC only], missing call rate >5%, and minor allele frequency (MAF) <5%.

We then performed analyses for five phenotype definitions: CLO, CPO, CLP, CL/P, and ANY.

### Genome-wide association analysis

For each phenotype, we analyzed the case-parent trios in GENEVA OFC with the transmission/disequilibrium test (TDT) as implemented in PLINK(41). For the POFC1 and POFC2 studies, we performed separate mixed logistic regression models using an additive genotype model as implemented in GENESIS, with a saddle point p-value approximation to account for case/control sample size imbalance and using a genetic relatedness matrix derived from PC-Relate and adjusting for 10 and 8 principal components of ancestry (PCAs), respectively, derived with PC-AiR(42,43). Results were combined across the three studies using a p-value-based meta-analysis with METAL(44). Regional association plots and credible sets for novel loci were generated using LocusZoom(45). We annotated variants and identified independent lead SNPs (those with meta-analysis p < 5e-8 and LD r^2^ < 0.1 with another lead SNP) among associated loci from the meta-analysis summary statistics with FUMA(46).

### Multi-tissue transcriptome-wide association analysis

Using S-PrediXcan(47), we combined cis-eQTL data from all tissues in the GTEx Project (version 8) (23) with our meta-analysis summary statistics using pre-specified weights based on covariance of genetic variants from PredictDB(48). We then combined S-PrediXcan data from all tissues and jointly analyzed them with S-MulTiXcan(49). We used a Bonferroni-adjusted significance threshold of 2.29e-6 (i.e., 0.05/21838) to account for the number of genes tested.

Gene set enrichment analyses of the significant genes identified in TWAS was performed using g:Profiler using a multiple query (separate gene lists for each phenotype definition) and using an adjusted p-value to maintain an experiment-wide type 1 error rate of 0.05 with the g:SCS algorithm that accounts for dependencies among gene annotations (11).

## Data availability statement

The data used in this study come from the International Consortium to Identify Genes and Interactions Controlling Oral Clefts, the Center for Craniofacial and Dental Genetics: Genetics of Orofacial Clefts and Related Phenotypes study, and the Center for Craniofacial and Dental Genetics: Genetics of Orofacial Clefts, Sub-types, and Subclinical Phenotypes study which are available in the database of Genotypes and Phenotypes (dbGaP) accession numbers phs000094.v1.p1, phs000774.v2.p1, and phs002815.v2.p1, respectively. In addition, Phenotypes, demographics, pregnancy history, and medical history available through FaceBase (facebase.org) for POFC1 (Site: https://www.facebase.org/chaise/record/#1/isa:dataset/RID=5A-FJBJ; Record ID: 5A-FJBJ; Accession #: FB00001369; DOI: <u>10.25550/5A-FJBJ) and POFC2</u> (Site: https://www.facebase.org/chaise/record/#1/isa:dataset/RID=56-ES6P; Record ID: 56-ES6P; Accession #: FB00001368; DOI: <u>10.25550/56-ES6P).</u>

## Data Availability

The data used in this study come from the International Consortium to Identify Genes and Interactions Controlling Oral Clefts, the Center for Craniofacial and Dental Genetics: Genetics of Orofacial Clefts and Related Phenotypes study, and the Center for Craniofacial and Dental Genetics: Genetics of Orofacial Clefts, Sub-types, and Subclinical Phenotypes study which are available in the database of Genotypes and Phenotypes (dbGaP) accession numbers phs000094.v1.p1, phs000774.v2.p1, and phs002815.v2.p1, respectively. In addition, Phenotypes, demographics, pregnancy history, and medical history available through FaceBase (facebase.org) for POFC1 (Site:  https://www.facebase.org/chaise/record/#1/isa:dataset/RID=5A-FJBJ; Record ID: 5A-FJBJ; Accession #: FB00001369; DOI: 10.25550/5A-FJBJ) and POFC2 (Site: https://www.facebase.org/chaise/record/#1/isa:dataset/RID=56-ES6P; Record ID: 56-ES6P; Accession #: FB00001368; DOI: 10.25550/56-ES6P).

## Acknowledgements

We thank the study participants for their participation and acknowledge the local recruitment staff and collaborators for their tireless efforts that made this study possible.

## Author contributions

**JCC**: Conceptualization, Formal Analysis, Methodology, Software, Supervision, Visualization, Writing – Original Draft Preparation; **XZ**: Formal Analysis, Methodology, Software, Visualization, Writing – Original Draft Preparation; **ZEY**: Formal Analysis, Software, Visualization, Writing – Original Draft Preparation, Writing – Review & Editing; **THB**: Data Curation, Writing – Review & Editing; **AB**: Data Curation, Funding Acquisition, Writing – Review & Editing; **CJB**: Data Curation, Writing – Review & Editing; **LJJG**: Data Curation, Writing – Review & Editing; **JTH**: Data Curation, Writing – Review & Editing; **RL**: Data Curation, Writing – Review & Editing; **LM**: Data Curation, Writing – Review & Editing; **JCM**: Data Curation, Funding Acquisition, Writing – Review & Editing; **IMO**: Data Curation, Writing – Review & Editing; **CP**: Data Curation, Writing – Review & Editing; **GLW**: Data Curation, Writing – Review & Editing; **EF**: Supervision, Writing – Review & Editing; **EJL**: Supervision, Writing – Review & Editing; **SMW**: Funding Acquisition, Project Administration, Resources, Supervision, Writing – Review & Editing; **MLM**: Data Curation, Funding Acquisition, Project Administration, Resources, Writing – Review & Editing; **JRS**: Conceptualization, Data Curation, Funding Acquisition, Project Administration, Resources, Supervision, Writing – Review & Editing

## Supporting Information Captions

**S1 Table. Association results for all genome-wide significant variants (p < 5e-8) from meta-analysis.** Columns include chromosome (Chr), position in hg38 (Pos), reference and alternate (effect) alleles (Ref/Alt), rsID, nearest gene, alternate allele frequency (AAF) for POFC1, POFC2, and GENEVA OFC, p-values and direction of effects for POFC1, POFC2, and GENEVA OFC, respectively (? indicates variant was not present in that study) for the five cleft phenotypes. Abbreviations Any: any cleft, CL/P: cleft lip with/without cleft palate, CLP: cleft lip and palate, CLO: cleft lip only, CPO: cleft palate only.

S2 Table. Results from multi-tissue TWAS for each gene surpassing the significance threshold (p < 2.29e-6) in at least one phenotype. Columns include Ensemble gene id, gene name, and p-values from the multi-tissue TWAS by each phenotype. Abbreviations: ANY = any cleft, CL/P = cleft lip with/without cleft palate, CLP = cleft lip and palate, CLO = isolated cleft lip, CPO = isolated cleft palate.

**S1 Figure. Manhattan and quantile-quantile plots of meta-analysis results from METAL for any cleft (ANY).** Plots were created with LocusZoom (45). Genomic control factor based on the median, λ_GC_, was 1.028.

S2 Figure. Manhattan and quantile-quantile plots of meta-analysis results from METAL for cleft lip with/without cleft palate (CL/P). Plots were created with LocusZoom (45). Genomic control factor based on the median, λ_GC_, was 1.024.

**S3 Figure. Manhattan and quantile-quantile plots of meta-analysis results from METAL for cleft lip and palate (CLP).** Plots were created with LocusZoom (45). Genomic control factor based on the median, λ_GC_, was 1.02.

**S4 Figure. Manhattan and quantile-quantile plots of meta-analysis results from METAL for cleft lip only (CLO).** Plots were created with LocusZoom (45). Genomic control factor based on the median, λ_GC_, was 0.99.

**S5 Figure. Manhattan and quantile-quantile plots of meta-analysis results from METAL for cleft palate only (CPO).** Plots were created with LocusZoom (45). Genomic control factor based on the median, λ_GC_, was 1.00.

**S6 Figure. Manhattan and quantile-quantile plots of multi-tissue TWAS results from S-MulTiXcan for any cleft (ANY).** Plots were created with LocusZoom (45) and qqman (50). Genomic control factor based on the median, λ_GC_, was 0.92.

S7 Figure. Manhattan and quantile-quantile plots of multi-tissue TWAS results from S-MulTiXcan cleft lip with/without cleft palate (CL/P). Plots were created with LocusZoom (45) and qqman (50). Genomic control factor based on the median, λ_GC_, was 0.941.

**S8 Figure. Manhattan and quantile-quantile plots of multi-tissue TWAS results from S-MulTiXcan for cleft lip and palate (CLP).** Plots were created with LocusZoom (45) and qqman (50). Genomic control factor based on the median, λ_GC_, was 0.917.

**S9 Figure. Manhattan and quantile-quantile plots of multi-tissue TWAS results from S-MulTiXcan for cleft lip only (CLO).** Plots were created with LocusZoom (45) and qqman (50). Genomic control factor based on the median, λ_GC_, was 0.842.

**S10 Figure. Manhattan and quantile-quantile plots of multi-tissue TWAS results from S-MulTiXcan for cleft palate only (CPO).** Plots were created with LocusZoom (45) and qqman (50). Genomic control factor based on the median, λ_GC_, was 0.856.

